# GENOTYPIC SPECTRUM OF ALBINISM IN MALI

**DOI:** 10.1101/2024.03.05.24303017

**Authors:** Modibo Diallo, Ousmane Sylla, Mohamed Kole Sidibé, Claudio Plaisant, Elina Mercier, Angèle Sequeira, Sophie Javerzat, Aziz Hadid, Eulalie Lasseaux, Vincent Michaud, Benoit Arveiler

## Abstract

Albinism is a phenotypically and genetically heterogeneous condition characterized by a variable degree of hypopigmentation and by ocular features leading to reduced visual acuity. Whereas numerous genotypic studies have been conducted throughout the world, very little is known about the genotypic spectrum of albinism in Africa and especially in Sub-Saharan Western Africa. Here we report the analysis of all 20 known albinism genes in a series a 23 patients originating from Mali. Four were diagnosed with OCA 1 (oculocutaneous albinism type 1), 17 with OCA 2, and 2 with OCA 4. *OCA2* variant NM_000275.3:c.819_822delinsGGTC was most frequently encountered. Four novel variants were identified (2 in *TYR*, 2 in *OCA2*). A deep intronic variant was found to alter splicing of the *OCA2* RNA by inclusion of a pseudo exon. Of note, the *OCA2* exon 7 deletion commonly found in eastern, central and southern Africa was absent from this series. African patients with OCA 1 and OCA 4 had only been reported twice and once respectively in previous publications. This study constitutes the first report of the genotypic spectrum of albinism in a western Sub-Saharan country.

## INTRODUCTION

Albinism is a genetic disease characterized by a variable degree of generalized hypopigmentation of the skin, hair and eye as well as by ocular features including nystagmus, misrouting at the optic chiasma, foveal hypoplasia and reduced visual acuity. Albinism is clinically and genetically heterogeneous with 20 genes involved in oculocutaneous, ocular and syndromic forms (for review see Bakker et al., 2022, Lasseaux et al., 2022). Worldwide frequency is difficult to estimate, especially because there are large differences between continents (Kromberg et al. 2023). In addition, while oculocutaneous albinism type 1 (OCA 1) is the most frequent form in Caucasian populations (Lasseaux et al, 2018), OCA 2 is most frequent in Africa (Kromberg and Kerr, 2022).

A 2.7Kb deletion encompassing exon 7 of the *OCA2* gene (Durham-Pierre et al., 1994) is highly prevalent in Eastern, Central and Southern Africa and originated 4,100-5,645 years in Bantu populations who migrated from their homeland at the Cameroon-Nigeria border towards the South following Eastern and Western routes that converged in Zimbabwe (Aquaron et al., 2007; Aquaron et al., 2019). OCA 1 seems to be very rare in Black Africans since only one patient was described (Badens et al., 2006), and another African American case was mentioned (King et al., 2003). OCA 3 in Black African patients was described as Rufous albinism, and is caused by African-specific variants of *TYRP1* (Manga et al., 1997). Only 1 Black African patient with OCA4 was described thus far (Moreno-Artero et al., 2022).

Only two studies analyzed the complete panel of the known albinism genes in African patients. One of them identified a patient from Senegal with Hermansky-Pudlak Syndrome type 1, a form of syndromic albinism (Ndiaye et al., 2019). In the second study, a patient from the Democratic Republic of Congo with both albinism and beta-thalassemia was found to have OCA 2 (Aquaron et al., 2022).

Here we analyzed a series of 23 patients from Mali. All presented clinically with oculocutaneous albinism. Sequencing of the 20 known albinism genes allowed establishing a molecular diagnosis in all patients, 4/23 with OCA 1, 17/23 with OCA 2, and 2/23 with OCA 4. We identified 4 new variants in *TYR*, *OCA2*. None of the patients harbored the *OCA2* exon 7 deletion.

## RESULTS

### Clinical findings in the patients

All 23 patients included in this study presented clinically with oculocutaneous albinism. They originated from various ethnic groups (Bambara, Forgeron, Malinke, Mianka, Peuhl, Sarakole, Senoufo, Soninke, Souraka), and were between 1 and 65 years old at the time of consultation. They all benefited from examination by both a dermatologist and an ophthalmologist at the Infirmerie Hôpital Militaire of Bamako (Mali). Skin and hair were severely hypopigmented. Skin color ranged from white to light brown, and hair color was white platinum or yellow in all patients except from patient 7 who had light brown hair. Nevi were present in patients 9 and 10 (65-69 and 25-29 year ranges, respectively). Ocular phenotype in all patients included nystagmus, various grades of foveal hypoplasia, iris transillumination, retinal hypopigmentation, and myopia (see Supplementary Figure 1 for an example of ocular phenotype). Visual acuity was between <1/10 and 3/10.

Main phenotypic traits of all patients are indicated in Table 1. See Supplementary Table 1 for a complete phenotypic description.

**Table 1:** Main phenotypic features of the 23 patients. F: female ; M: male ; NS: not shown ; NA: Not available. VA RE/LE: Visual acuity Right Eye/Left Eye. ITI: Iris transillumination. RHP: Retinal hypopigmentation. FHP: Foveal hypoplasia (Thomas et al., 2011 classification).

338G>T; p.(Glu280Ter). Transcript used of each gene: *TYR* NM\_000372.5; *OCA2* NM\_000275.3; *SLC45A2* NM\_016180.5.
| Patient | Gender | Age range in years | Consanguinity | Skin color | Hair color | Iris color | Nystagmus | Amblyopia |
| --- | --- | --- | --- | --- | --- | --- | --- | --- |
| 1 | F | 5-9 | Yes | White | Yellow | Light blue | Yes | Myopia |
| 2 | M | 10-14 | No | White | Yellow | Brown | Yes | Myopia |
| 3 (sibling of patient 2) | F | 10-14 | No | White | Yellow | Brown | Yes | Myopia |
| 4 | F | 5-9 | No | White | Yellow | Brown | Yes | Myopia |
| 5 | F | 0-4 | Yes | Light brown | Yellow | Blue | Yes | Myopia |
| 6 | M | 30-34 | Yes | White | Yellow | Brown | Yes | NA |
| 7 | F | 10-14 | No | Light brown | Brown light | Brown | Yes | Myopia |
| 8 | F | 10-14 | No | Light brown | Yellow | Brown | Yes | Myopia |
| 9 | NS | 65-69 | Yes | White | Yellow | Brown | Yes | Myopia |
| 10 | M | 25-29 | No | White | Yellow | Brown | Yes | Myopia |
| 11 | M | 5-9 | No | Light brown | Yellow | Brown | Yes | Myopia |
| 12 | F | 10-14 | No | Light brown | Yellow | Brown | Yes | NA |
| 13 | M | 10-14 | No | Light brown | Yellow | Brown | Yes | Myopia |
| 14 | F | 10-14 | Yes | White | Yellow | Brown | Yes | Myopia |
| 15 | F | 15-19 | Yes | White | Yellow | Brown | Yes | Myopia |
| 16 | F | 0-4 | Yes | Pink | Yellow | Brown | Yes | Myopia |
| 17 | M | 30-34 | Yes | White | Yellow | Brown | Yes | NA |
| 18 | M | 20-24 | Yes | White | NA | Brown | Yes | Myopia |
| 19 | M | 10-14 | Yes | Light brown | Yellow | Brown | Yes | Myopia |
| 20 (sibling of patient 19) | M | 10-14 | Yes | White | Yellow | Brown | Yes | Myopia |
| 21 | M | 15-19 | Yes | White | Yellow | Brown | Yes | Myopia |
| 22 | NS | 30-35 | Yes | White | Platine white | Brown | Yes | Myopia |
| 23 | F | 0-4 | Yes | Light brown | Yellow | Brown | Yes | Myopia |

| VA RE/LE | ITI | RHP | FHP |
| --- | --- | --- | --- |
| RE<1/10 LE<1/10 | Stage 3 | Yes | Stage 3 |
| RE<1/10 LE<1/10 | Stage 3 | Stage 3 | Yes |
| RE<1/10 LE<1/10 | Stage 3 | Stage 3 | Yes |
| RE<1/10 LE<1/10 | Stage 3 | Stage 4 | Stage 4 |
| NA | Stage 3 | Stage 3 | NA |
| NA | NA | NA | NA |
| RE=2/10 LE= 1/10 | Stage 3 | Yes | Stage 2 |
| RE=2/10 LE=2/10 | Stage 3 | Stage 3 | NR |
| RE<1/10 LE<1/10 | Stage 3 | Stage 4 | Stage 4 |
| RE=1/10 LE=3/10 | Stage 3 | Yes | Stage 3 |
| RE<1/10 LE=3/10 | Stage 2 | Stade 3 | Stage 3 |
| RE<1/10 LE <1/10 | Stage 3 | Stade 2 | Stage 3 |
| RE<1/10 LE<1/10 | Yes | Yes | Yes |
| RE<1/10 LE<1/10 | NA | Stage 4 | Stage 4 |
| RE<1/10 LE<1/10 | Stage 3 | Stage 3 | Stage 3 |
| NA | Stage 3 | Stage 3 | Stage 2 |
| NA | NA | NA | NA |
| RE<1/10 LE=1/10 | Stage 3 | Yes | Stage 2 |
| RE<1/10 LE<1/10 | Stage 3 | Stage 4 | Stage 4 |
| RE<1/10 LE<1/10 | Stage 4 | Stage 4 | Stage 4 |
| RE<1/10 LE=2/10 | Stage 3 | Stage 4 | Stage 3 |
| RE<1/10 LE<1/10 | Stage 3 | Stage 4 | Stage 4 |
| NA | Stage 3 | Stage 4 | Stage 3 |

### Genotypic spectrum

Sequencing of the 20 known albinism genes allowed to establish a molecular diagnosis in all 23 patients, based upon the identification of 2 class 4 (likely pathogenic) or 5 (pathogenic) variants according to the American College of Medical Genetics (ACMG) criteria (Richards et al., 2015). Parental segregation of variants was established for all patients, except for the elder ones (9 and 22) whose parents were not available for analysis, and was consistent with autosomal recessive inheritance with compound heterozygous or homozygous variants. Four patients were diagnosed with OCA 1, 17 with OCA 2, and 2 with OCA 4. Variants identified are indicated for each patient in Table 2 and Supplementary Table 1. ACMG classification criteria are not commented in the following paragraphs for already published variants, but are presented for the new variants (ClinVar Submission ID: SUB13987935).

**Table 2.** Genotypes of the 23 patients. Novel variants are in Bold. $: Patient 23 had also a heterozygous variant in *TYR* NM_000372.5:c.838G>T; p.(Glu280Ter). Transcript used of each gene: *TYR* NM_000372.5; *OCA2* NM_000275.3; *SLC45A2* NM_016180.5.

| Patient | Consanguinity | Gene | Variant 1 | Localisation | ACMG |
| --- | --- | --- | --- | --- | --- |
| 1 | Yes | *TYR* | **c.255del; p.(Tyr85\*)** | Exon 1 | Class 5 |
| 2 | No | *TYR* | **c.259A>G; p.(Arg87Gly)** | Exon 1 | Class 4 |
| 3 (sibling of patient 2) | No | *TYR* | **c.259A>G; p.(Arg87Gly)** | Exon 1 | Class 4 |
| 4 | No | *TYR* | c.880G>A; p.(Glu294Lys) | Exon 2 | Class 4 |
| 5 | Yes | *OCA2* | c.819_822delinsGGTC ;p.(Asn273_Trp274delinsLysVal) | Exon 8 | Class 5 |
| 6 | Yes | *OCA2* | c.819_822delinsGGTC ;p.(Asn273_Trp274delinsLysVal) | Exon 8 | Class 5 |
| 7 | No | *OCA2* | c.819_822delinsGGTC ;p.(Asn273_Trp274delinsLysVal) | Exon 8 | Class 5 |
| 8 | No | *OCA2* | c.819_822delinsGGTC ;p.(Asn273_Trp274delinsLysVal) | Exon 8 | Class 5 |
| 9 | Yes | *OCA2* | c.819_822delinsGGTC ;p.(Asn273_Trp274delinsLysVal) | Exon 8 | Class 5 |
| 10 | No | *OCA2* | c.819_822delinsGGTC ;p.(Asn273_Trp274delinsLysVal) | Exon 8 | Class 5 |
| 11 | No | *OCA2* | c.819_822delinsGGTC ;p.(Asn273_Trp274delinsLysVal) | Exon 8 | Class 5 |
| 12 | No | *OCA2* | c.819_822delinsGGTC ;p.(Asn273_Trp274delinsLysVal) | Exon 8 | Class 5 |
| 13 | No | *OCA2* | c.819_822delinsGGTC ;p.(Asn273_Trp274delinsLysVal) | Exon 8 | Class 5 |
| 14 | Yes | *OCA2* | c.1349C>T; p.(Thr450Met) | Exon 13 | Class 4 |
| 15 | Yes | *OCA2* | c.1349C>T; p.(Thr450Met) | Exon 13 | Class 4 |
| 16 | Yes | *OCA2* | c.2339G>A; p.(Gly780Asp) | Exon 23 | Class 5 |
| 17 | Yes | *OCA2* | c.2339G>A; p.(Gly780Asp) | Exon 23 | Class 5 |
| 18 | Yes | *OCA2* | c.2339G>A; p.(Gly780Asp) | Exon 23 | Class 5 |
| 19 | Yes | *OCA2* | c.2339G>A; p.(Gly780Asp) | Exon 23 | Class 5 |
| 20 (twin of patient 19) | Yes | *OCA2* | c.2339G>A; p.(Gly780Asp) | Exon 23 | Class 5 |
| 21 | Yes | *OCA2* | c.2339G>A; p.(Gly780Asp) | Exon 23 | Class 5 |
| 22 | Yes | *SLC45A2* | c.977T>A; p.(Ile326Asn) | Exon 4 | Class 4 |
| 23 | Yes | *SLC45A2*\$ | c.977T>A; p.(Ile326Asn) | Exon 4 | Class 4 |

| Criteria | Variant 2 | Localisation | ACMG | Criteria |
| --- | --- | --- | --- | --- |
| PVS1 PM2 | **c.255del; p.(Tyr85\*)** | Exon 1 | Class 5 | PVS1 PM2 |
| PM1 PM2 PP1 PP3 | **c.259A>G; p.(Arg87Gly)** | Exon 1 | Class 4 | PM1 PM2 PP1 PP3 |
| PM1 PM2 PP1 PP3 | **c.259A>G; p.(Arg87Gly)** | Exon 1 | Class 4 | PM1 PM2 PP1 PP3 |
| PM1 PM2 PP3 PP5 | c.1115G>A; p.(Gly372Glu) | Exon 3 | Class 4 | PM1 PM2 PP3 PP5 |
| PS4 PM1 PM2 PP3 PP5 | c.819_822delinsGGTC ;p.(Asn273_Trp274delinsLysVal) | Exon 8 | Class 5 | PS4 PM1 PM2 PP3 PP5 |
| PS4 PM1 PM2 PP3 PP5 | c.819_822delinsGGTC ;p.(Asn273_Trp274delinsLysVal) | Exon 8 | Class 5 | PS4 PM1 PM2 PP3 PP5 |
| PS4 PM1 PM2 PP3 PP5 | c.819_822delinsGGTC ;p.(Asn273_Trp274delinsLysVal) | Exon 8 | Class 5 | PS4 PM1 PM2 PP3 PP5 |
| PS4 PM1 PM2 PP3 PP5 | c.819_822delinsGGTC ;p.(Asn273_Trp274delinsLysVal) | Exon 8 | Class 5 | PS4 PM1 PM2 PP3 PP5 |
| PS4 PM1 PM2 PP3 PP5 | c.819_822delinsGGTC ;p.(Asn273_Trp274delinsLysVal) | Exon 8 | Class 5 | PS4 PM1 PM2 PP3 PP5 |
| PS4 PM1 PM2 PP3 PP5 | **c.759del; p.(Glu253Aspfs\*2)** | Exon 7 | Class 5 | PVS1 PM2 |
| PS4 PM1 PM2 PP3 PP5 | c.2339G>A; p.(Gly780Asp) | Exon 23 | Class 4 | PM1 PM2 PP3 PP5 |
| PS4 PM1 PM2 PP3 PP5 | c.2378G>A; p.(Cys793Tyr) | Exon 23 | Class 5 | PM1 PM2 PM5 PP3 |
| PS4 PM1 PM2 PP3 PP5 | c.2425T>A; p.(Phe809Ile) | Exon 23 | Class 4 | PM1 PM2 PP3 PP5 |
| PM1 PM2 PP3 PP5 | c.1349C>T; p.(Thr450Met) | Exon 13 | Class 4 | PM1 PM2 PP3 PP5 |
| PM1 PM2 PP3 PP5 | c.1349C>T; p.(Thr450Met) | Exon 13 | Class 4 | PM1 PM2 PP3 PP5 |
| PM1 PM2 PP3 PP5 | c.2339G>A; p.(Gly780Asp) | Exon 23 | Class 5 | PM1 PM2 PP3 PP5 |
| PM1 PM2 PP3 PP5 | c.2339G>A; p.(Gly780Asp) | Exon 23 | Class 5 | PM1 PM2 PP3 PP5 |
| PM1 PM2 PP3 PP5 | c.2339G>A; p.(Gly780Asp) | Exon 23 | Class 5 | PM1 PM2 PP3 PP5 |
| PM1 PM2 PP3 PP5 | **c.1951+1215G>T; p.(Gly651_Trp652ins\*18)** | Intron 18 | Class 5 | PVS1 PS3 PM2 PM3 |
| PM1 PM2 PP3 PP5 | **c.1951+1215G>T; p.(Gly651_Trp652ins\*18)** | Intron 18 | Class 5 | PVS1 PS3 PM2 PM3 |
| PM1 PM2 PP3 PP5 | c.2425T>A; p.(Phe809Ile) | Exon 23 | Class 5 | PM1 PM2 PP3 PP5 |
| PS4 PM1 PM2 PP3 | c.977T>A; p.(Ile326Asn) | Exon 4 | Class 4 | PS4 PM1 PM2 PP3 |
| PS4 PM1 PM2 PP3 | c.977T>A; p.(Ile326Asn) | Exon 4 | Class 4 | PS4 PM1 PM2 PP3 |

### TYR (OCA1) variants

Two known variants, NM_000372.5:c.880G>A; p.(Glu294Lys) and NM_000372.5:c.1115G>A; p.(Gly372Glu), were found in the compound heterozygous state in patient 4.

Two new *TYR* variants are presented hereafter.

NM_000372.5:c.255del; p.(Tyr85*) was found in the homozygous state in patient 1. This nonsense variant, absent from GnomAD (GnomADv3.2 https://gnomad.broadinstitute.org/), was rated PVS1 PM2 (pathogenic, class 5).

NM_000372.5:c.259A>G; p.(Arg87Gly) was found in the homozygous state in patients 2 and 3, who are siblings. This variant is absent from the control population database GnomAD. Arg87 is highly conserved in 10/12 species and in the TYR/TYRP1/TYRP2(DCT) family of proteins in humans. The physicochemical difference between Arg and Gly is moderate (Grantham score: 125). This variant was rated PM1 PM2 PP1 PP3 (probably pathogenic, class 4) according to ACMG criteria (Richards et al., 2015).

### OCA2 variants

Nine patients molecularly diagnosed with OCA 2 carried the NM_000275.3:c.819_822delinsGGTC; p.(Asn273_Trp274delinsLysVal) class 5 variant (Lee et al. 1994), 5/9 and 4/9 in the homozygous and compound heterozygous states, respectively.

The second most frequent *OCA2* variant in our series, NM_000275.3:c.2339G>A; p.(Gly780Asp) (class 5) was present in 3 patients in the homozygous state and in 4 in the compound heterozygous state.

NM_000275.3:c.1349C>T; p.(Thr450Met) (class4) was present in 2 siblings (homozygous), NM_000275.3:c.2425T>A; p.(Phe809Ile) (class 5) in 2 patients (compound heterozygous), and NM_000275.3:c.2378G>A; p.(Cys793Tyr) (class 4) in 1 patient (compound heterozygous).

Two new *OCA2* variants were identified, both in the compound heterozygous state with another class 5 variant (patients 10, 19 and 20).

NM_000275.3:c.759del; p.(Glu253Aspfs*2) is a frameshift variant (class 5, PVS1 PM2), absent from gnomADv3.2.

NM_000275.3:c.1951+1215G>T (g.27950569C>A), located deep in *OCA2* intron 18, was identified in the compound heterozygous state in two siblings (patients 19 and 20). This is a very rare variant (2 heterozygotes, 0 homozygote in gnomADv3.2). Bioinformatic predictions using the RNA-Splicer software (https://rddc.tsinghua-gd.org/) and SpliceAI-visual integrated in the Mobidetails variant interpretation tool (https://mobidetails.iurc.montp.inserm.fr/MD) suggested that this variant could activate a cryptic acceptor splice site at coordinate c.1951+1219 and a cryptic donor splice site at c.1951+1297, thereby including a 77 bp pseudo-exon (g.27950564-27950488) (Supplementary Figure 2). We recently showed that *OCA2* is expressed in blood cells at a level compatible with RT-PCR analysis (Michaud et al., 2023). Total RNA was extracted from a blood sample of patient 20 and RT-PCR was used to amplify the transcribed sequences between exons 16 and 23. A 648 bp PCR product corresponding to normal splicing was observed as expected in a control individual not harboring the variant. Instead a specific 725 bp PCR product was obtained in the patient (Figure 1). Sanger sequencing showed that this product included the 77 bp pseudoexon. It can be noted that the second (normal) allele was not amplified under the RT-PCR conditions used (annealing of primers at 65°C) but was amplified under less stringent conditions (annealing at 63°) (not shown). Hence variant NM_000275.3:c.1951+1215G>T triggers the inclusion of a 77 bp pseudoexon in the *OCA2* RNA (r.1951_1952ins1951+1220_1951+1296). Translation of the pseudoexon is predicted to hit a premature stop codon after 17 amino acids, thus producing a truncated P protein (NP_000266.2:p.(Gly651_Trp652ins*18). This variant is therefore classified as pathogenic (PVS1 PS3 PM2 PM3), allowing to establish the OCA 2 diagnosis in patients 19 and 20.

**Figure 1:**
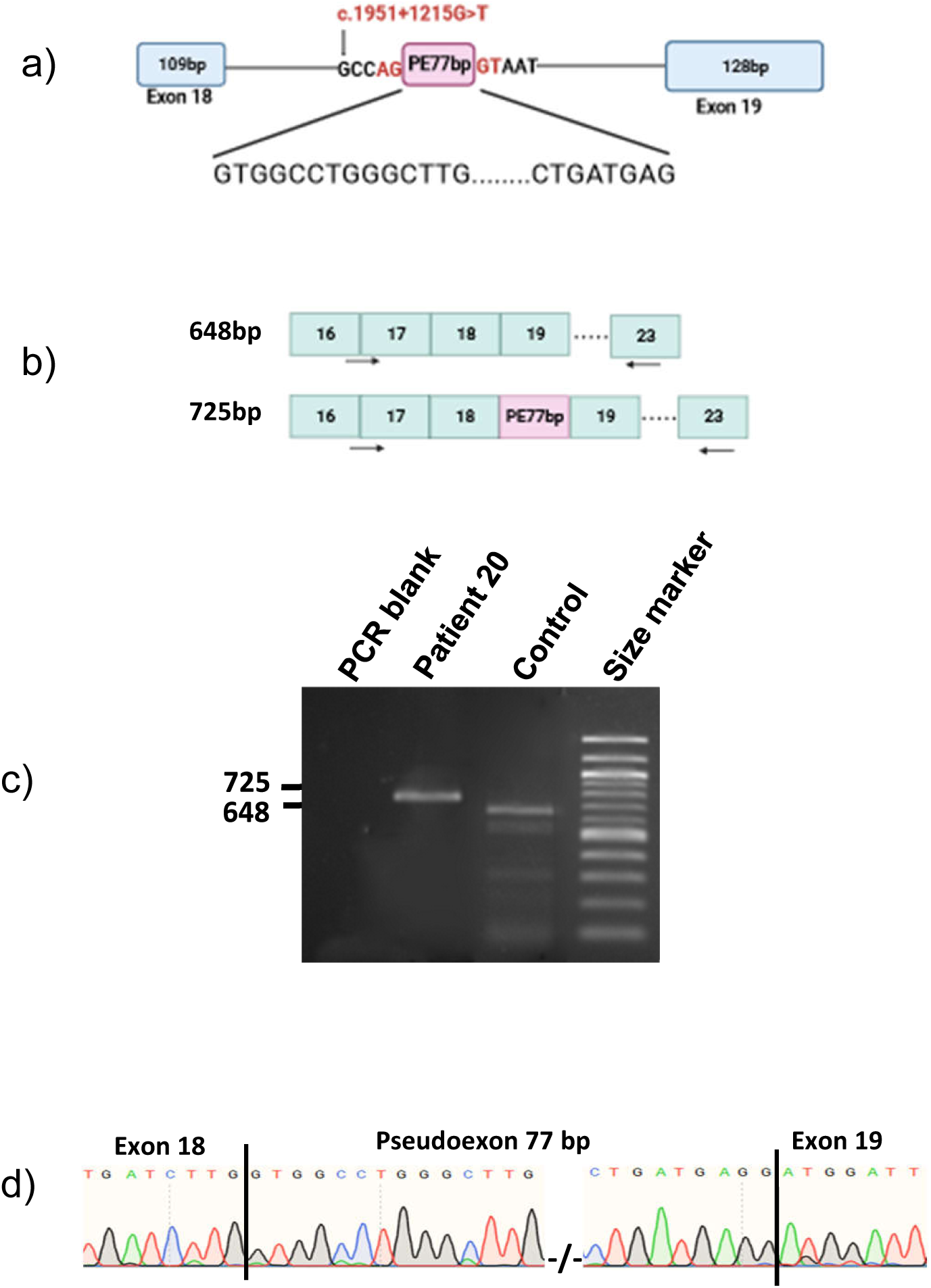
RT-PCR analysis of *OCA2* intronic variant c.1951+1215G>T in patient 20. a) Schematic view of the genomic region encompassing exon 18, intron 18 and exon 19. The deep intronic variant c.1951+1215G>T is shown in red. The 77 bp pseudoexon (PE) is displayed as a pink box. Intronic sequences surrounding the pseudoexon are indicated on the genome-representing line. Sequences at the beginning and end of the pseudo exon are shown underneath. b) Design of the RT-PCR assay showing the expected sizes for the RT-PCR products without (648 bp) and with (725 bp) the pseudoexon. Exons are represented as blue boxes with exon numbers indicated. The pseudoexon (PE) is in pink. RT-PCR Primers are shown as black arrows. c) Agarose gel electrophoresis of the RT-PCR products from blood cells. Patient 20 shows a band at 725 bp expected to include the pseudoexon. The control individual shows the band at 648 bp expected not to include the pseudoexon. Size marker is a 100 bp DNA ladder. d) Sanger sequencing electrophoregram of the 725bp RT-PCR product from patient 20 showing that the 77 bp pseudoexon is included between *OCA2* exons 18 and 19.

### SLC45A2 (OCA4) variants

Two unrelated patients (22 and 23) were diagnosed with OCA 4. Both were homozygous for a variant of *SLC45A2*, NM_016180.5:c.977T>A;p.(Ile326Asn), a very rare variant (3 heterozygotes, 0 homozygote in gnomADv3.2) that was recently described by Moreno et al., 2022 in a French patient. Ile326 is moderately conserved (present in 8/12 species) and there is a large physicochemical difference between Ile and Asn (Grantham score dis =149 (0-215). We rated this variant PS4 PM1 PM2 PP3 (class 4).

Of note, patient 23 also had a new nonsense *TYR* variant NM_000372.5:c.838G>T; p.(Glu280Ter) (PVS1 PM2, class 5). Despite sequencing of the entire *TYR* gene (see Materials and Methods), no other pathogenic variant could be identified, thus indicating that the patient does not have OCA 1 in addition to OCA 4.

## DISCUSSION

We genotyped 23 patients with a typical oculocutaneous albinism phenotype from Mali. A molecular diagnosis was obtained in all of them. Four patients had OCA 1 (17.4%), 17 had OCA 2 (73.9%), and 2 had OCA 4 (8.7%).

Only 1 African patient had been described so far with OCA 1 (Badens et al., 2006), and another one mentioned in a larger series (King et al., 2003). It is therefore remarkable that we identified 4 new OCA 1 patients. One was compound heterozygous for already described pathogenic variants, 2 siblings were homozygous for the new variant NM_000372.5:c.259A>G; p.(Arg87Gly) and 1 was homozygous for the new variant NM_000372.5:c.255del; p.(Tyr85*). Of note, patient 23 otherwise diagnosed with OCA 4 was heterozygous for an additional novel *TYR* pathogenic variant, NM_000372.5:c.838G>T; p.(Glu280Ter). Altogether, these results strongly suggest that more OCA 1 cases can be expected in this country and potentially in other Sub-Saharan countries.

Amongst patients with OCA 2, 9 carried the NM_000275.3:c.819_822delinsGGTC; p.(Asn273_Trp274delinsLysVal) variant either in the homozygous (5/9) or in the compound heterozygous state with another *OCA2* class 4 or 5 variant (4/9). This variant seems restricted to Black African patients, based on the publication by Lee et al. (1994) and the fact that all 31 patients from our cohort of more than 2000 patients for whom geographical origin was documented in the clinical record were Black Africans. These include cases already published by us (Lasseaux et al., 2018; Marti et al., 2018) and unpublished ones. Of note the NM_000275.3:c.2425T>A; p.(Phe809Ile) variant present in 2 patients also seems African-specific since the 10 patients from our cohort with geographical origin documented who harbored it were all of Sub-Saharan origin (unpublished data).

We identified 2 new *OCA2* variants, NM_000275.3:c.759del; p.(Glu253Aspfs*2) and NM_000275.3:c.1951+1215G>T; p.Gly651_Trp652ins*18. The latter alters splicing of the *OCA2* mRNA with the inclusion of an intron 18-derived pseudoexon as assessed by RT-PCR. Strikingly, the *OCA2* exon 7 deletion commonly encountered in Black African patients was absent in our series of patients, suggesting that this deletion is uncommon in Sub-Saharan Western Africa, fitting with the migration route of this deletion with the Bantu population from the Cameroon-Nigeria border towards the South of the continent (Aquaron et al., 2007; Aquaron et al., 2018).

Two unrelated patients were homozygous for the same *SLC45A2* variant NM_016180.5:c.977T>A; p.(Ile326Asn) (OCA 4). Interestingly both were from the same ethnic group.

Apart from this possible exemplar, no specific variant or OCA type could be consigned to a particular ethnic group. For instance, *OCA2* variant NM_000275.3:c.819_822delinsGGTC; p.(Asn273_Trp274delinsLysVal) was present in patients from 5 different ethnic groups, NM_000275.3:c.2339G>A; p.(Gly780Asp) as well, and NM_000275.3:c.2425T>A; p.(Phe809Ile) was present in patients from 2 different ethnic groups. In addition, Bambara patients had either OCA 1 or OCA 2. Information about ethnic groups can be obtained from the authors upon request.

From a phenotypical point of view, all patients displayed a typical and severe OCA phenotype with light skin and hair color, iris transillumination (100% of evaluated cases, grade 2-4), retinal hypopigmentation (100% of evaluated cases, grade 2-4), nystagmus (100% of cases), foveal hypoplasia (100% of evaluated cases, grade 2-4), myopia (100% of evaluated cases), and low visual acuity (<0.1 in the majority of cases, 0.2 or 0.3 in 4 cases) (see Table 1).

All patients diagnosed with OCA 1 had severe cutaneous and ocular phenotypes, whatever the type of variant (nonsense or missense).

Concerning patients with OCA 2, those homozygous for the NM_000275.3:c.819_822delinsGGTC; p.(Asn273_Trp274delinsLysVal) had light brown skin and yellow or light brown hair, indicating the existence of some degree of pigmentation, except from patient 9 who had white skin and yellow hair. Those who were compound heterozygous for this DelIns variant and a missense variant had all yellow hair, and light brown skin. Patient 10 was compound heterozygous with a frameshift variant and had white skin and yellow hair. There was no salient difference at the ocular level between homozygous and compound heterozygous patients. The 2 patients homozygous for NM_000275.3:c.1349C>T; p.(Thr450Met) and those either homozygous or compound heterozygous for variant NM_000275.3:c.2339G>A; p.(Gly780Asp) had severe oculocutaneous albinism, including the patients harboring the novel deep intronic variant NM_000275.3:c.1951+1215G>T; p.Gly651_Trp652ins*18.

Both patients with OCA 4 have severe hypopigmentation of the skin and hair as well as a severe ocular phenotype with grade 3 iris transillumination, grade 4 retinal hypopigmentation, and grade 3 or 4 foveal hypoplasia.

Interestingly, patients 9 and 22 are the parents of 3 clinically unaffected children (see Supplementary Figure 3). One parent (patient 9) was homozygous for *OCA2* variant NM_000275.3:c.819_822delinsGGTC; p.(Asn273_Trp274delinsLysVal) and had OCA 2, whereas the other (patient 22) was homozygous for *SLC45A2* variant NM_016180.5:c.977T>A; p.(Ile326Asn) and had OCA 4. The children were double heterozygotes for the parental variants. They had black skin although the level of pigmentation seemed slightly decreased compared to the rest of the Malian population. Careful examination indicated that one of the children had somewhat red hair whereas the other two had grey hair. Grade 1 hypopigmentation of the iris was observed in all three children, and one of them had red pupillary reflex (data not shown). The *OCA2*/*SLC45A2* double heterozygous progeny do not have albinism, but their mild oculocutaneous features suggest some degree of epistasis between the two genes that are both known to control melanosomal pH (Bellono et al., 2014; Le et al., 2020).

In conclusion, we describe the phenotypes and genotypes of 23 patients originating from Mali. Molecular diagnosis was obtained for all of them. Interestingly, 4 patients had OCA 1, whereas only 2 OCA 1 Black African patients) had been described so far, one from Cameroon (Badens et al., 2006), and one described as African American (King et al., 2003). In addition, we report OCA 4 in Sub-Saharan Africa for the second time, after the recent description of one Mauritanian patient (Moreno-Artero et al., 2022). Of note, 4 novel variants were identified in the *TYR* and *OCA2* genes. On the other hand, the common exon 7 deletion of the *OCA2* gene (Durham-Pierre et al., 1994) was absent from this series of patients. None of the patients had a syndromic form of OCA, while 1 patient from Senegal has been formerly reported with HPS1 (Ndiaye et al., 2019).

Publications about albinism in Africa were so far almost exclusively dedicated to patients from Central and Southern Africa (see Kromberg and Kerr, 2022 for review). Our study presents the first description of a series of patients with albinism originating from Western Sub-Saharan Africa, and the first study analyzing the complete set of albinism genes in a series of African patients. It will be interesting to extend this work to other patients from Mali and other Western Sub-Saharan countries in order to further documenting the genotypic spectrum of the disease in this part of the continent.

## MATERIALS AND METHODS

### Patients

All patients originated from Mali. Informed consent for genetic analysis and for taking part in a research study, as well as authorization for publication, including photographs, were obtained from the patients or their parents if minors. This study was approved by the ethics committee of the Faculty of Medicine of the University of Sciences, Techniques and Technologies of Bamako, Mali (USTTB).

### Next generation sequencing of the panel of albinism genes

The 20 known albinism genes were analyzed (*TYR, OCA2, TYRP1, SLC45A2, SLC24A5, LRMDA, TYRP2/DCT, GPR143, HPS1-11, CHS1*) (Bakker et al., 2022; Lasseaux et al., 2022). This included for all genes the exons and intron-exon junctions. The introns and flanking sequences were also analyzed for *TYR* (OCA 1), *OCA2* (OCA 2), *SLC45A2* (OCA 4), *GPR143* (OA 1), and *BLOC3S1* (HPS 1). Coordinates of all sequences included are available upon request.

Library preparation, capture, enrichment and elution were performed according to the manufacturer’s protocol (SureSelect XT HS Custom; Agilent Technologies). Each sample was sequenced in 75 bp paired-end reads on an Illumina NextSeq550Dx sequencer (Thermo Fisher Scientific). Alignment on the reference sequence (GRCh38) and variant calling (Single Nucleotide Variants and Copy Number Variants) were performed with Alissa Reporter (Agilent Technologies). Annotation and filtering of the variants were carried out with Alissa Interpret (Agilent Technologies). The sequence of the selected variants was visualized using Alamut Visual Plus (Sophia Genetics). A sample quality data check was performed. Details concerning the analytical method, bioinformatics analysis and versions of the tools and database used are available on request. Segregation analysis of the variants in the parents was performed by Sanger sequencing (BDT v3.1 on ABI3500xL Dx, Thermo Fisher Scientific). Pathogenicity prediction algorithms were implemented for each variant, including CADD, MPA score, MaxEntScan, SPiP, and SpliceAI-visual, integrated in MobiDetails (https://mobidetails.iurc.montp.inserm.fr/MD) (Baux et al., 2021), Alamut visual Plus (Sophia Genetics) and RNA-Splicer (https://rddc.tsinghua-gd.org/). The Minor allele frequency (MAF) was defined using the Genome Aggregation Database (gnomADv3.1.2) (https://gnomad.broadinstitute.org/) with a threshold ≤ 0,001.

The new variants identified in this study were deposited in ClinVar (Submission ID: SUB13987935).

### RT-PCR on blood samples

Total RNA was isolated from white blood cells obtained from patient 20, and from a control individual without any known genetic disease, using the PAXgene Blood RNA kit (Qiagen), as indicated in the manufacturer’s protocol. One µg of total RNA was reverse transcribed into cDNA using a cDNA synthesis Kit (Thermo Fisher Scientific). Reverse Transcription-PCR primers were designed (primer3 version 4.1.0; https://primer3.ut.ee/) based on the *OCA2* mRNA sequence (NM_000275.3). Forward primer (5’ GCACACCTTCCACAGACAGA 3’) was at the junction between exons 16 and 17, and reverse primer (5’AAGGAGAACCCATATCCATGC 3’) was in exon 23. PCR conditions were 40 cycles (95°for 30s, 65° for 15s and 72° for 20s). PCR products were separated by 2% agarose gel electrophoresis and Sanger sequenced (Eurofins).

## DATA AVAILABILITY STATEMENT

BED files indicating coordinates of the albinism NGS panel used for this study are available from the authors upon request.

The new variants identified in this study were deposited in ClinVar (Submission ID: SUB13987935).

## ACKNOWLEDGMENTS

The authors are grateful to the Association Malienne pour la protection des patients avec albinisme (AMPA) de Bamako (Mali) for their strong support, to the patients and their families for participating in the study, and to Lassana Sylla, Ibrahim Haïdara, Elodie Philippe, and Isabelle Helot for their excellent technical support. MD, OS and MKS are grateful to the Ministère de la Défense et des Anciens Combattants, the Etat-Major Général des Armées, the Direction Centrale des Services de Santé des Armées (DCSSA) and the Infirmerie Hôpital de Bamako (IHB) for authorizing this study and for the mobilization of staff. MD and BA are grateful to the Programme de Formation des Formateurs (PFF) du Ministère de l’Enseignement Supérieur et de la Recherche Scientifique du Mali for financing patients sequencing costs, and to Genespoir, the French albinism association, for their financial support to our research activities.

## AUTHOR CONTRIBUTIONS STATEMENT

Conceptualization: Modibo Diallo (lead), Ousmane Sylla (lead), Mohamed Kole Sidibé (lead), Benoit Arveiler (lead)

Funding Acquisition: Benoit Arveiler (lead), Modibo Diallo (lead)

Investigation: Modibo Diallo (lead), Ousmane Sylla (lead), Mohamed Kole Sidibé (lead), Claudio Plaisant (equal), Elina Mercier (equal), Angèle Sequeira (equal)

Methodology: Modibo Diallo (lead), Ousmane Sylla (lead), Mohamed Kole Sidibé (lead), Claudio Plaisant (equal), Elina Mercier (equal), Angèle Sequeira (equal)

Project Administration: Benoit Arveiler (lead)

Resources: Ousmane Sylla (lead), Mohamed Kole Sidibé (lead), Aziz Hadid (equal)

Supervision: Benoit Arveiler (lead)

Validation: Modibo Diallo (lead), Ousmane Sylla (equal), Mohamed Kole Sidibé (equal), Claudio Plaisant (equal), Elina Mercier, Angèle Sequeira, Benoit Arveiler (lead)

Visualization: Modibo Diallo (lead), Benoit Arveiler (lead)

Writing - Original Draft Preparation: Modibo Diallo (lead), Benoit Arveiler (lead)

Writing - Review and Editing: Ousmane Sylla (equal), Mohamed Kole Sidibé (equal), Sophie Javerzat (equal), Eulalie Lasseaux (equal), Vincent Michaud (equal), Benoit Arveiler (equal)

## ABBREVIATIONS USED

OCA: oculocutaneous albinism
ACMG: American College of Medical Genetics
bp: base pairs

