## Supplementary Figure 1 for "GENOTYPIC SPECTRUM OF ALBINISM IN MALI"

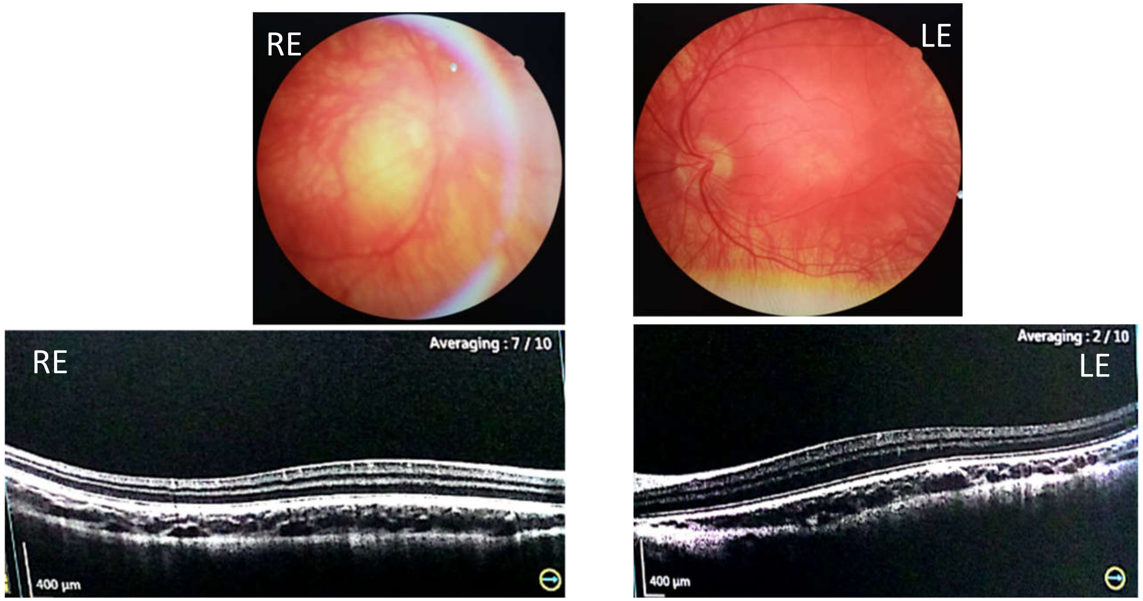


**Supplementary Figure 1: Ophthalmologic phenotype of Patient 13 with OCA 2 (c.[819_822delinsGGTC];[2425T>A]).** Eye fundi examination showed retinal hypopigmentation of right eye (RE) and left eye (LE) (upper panel). Optical Coherence Tomography (OCT) showing the foveal hypoplasia in right eye (RE) and left eye (LE) (lower panel). Informed consent for publication, including photographs, was obtained from the patients or their parents if minors.
