## Supplementary Figure 2 for "GENOTYPIC SPECTRUM OF ALBINISM IN MALI"

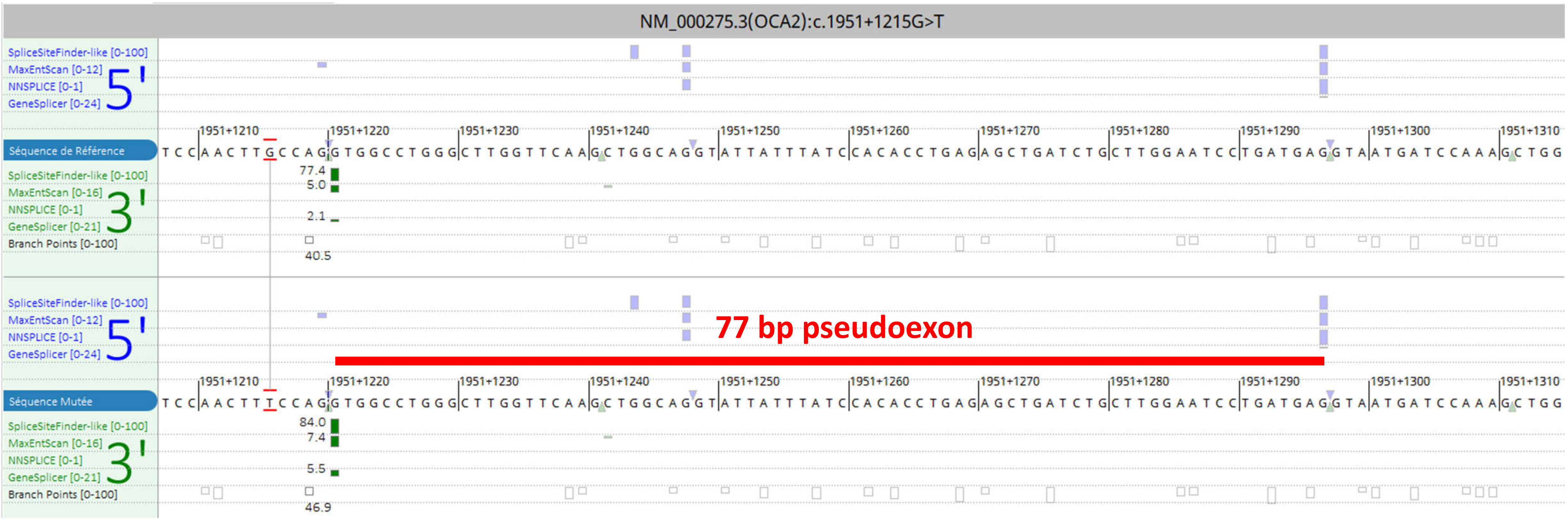


**Supplementary Figure 2A: ALAMUT VISUAL PLUS PREDICTION.**

Alamut Visual Plus (Agilent Technologies) shows the genomic region around the NM_000275.3:c.1951+1215G>T variant. The wild type G (upper panel) and variant T (lower panel) nucleotides are between red lines. The crytpic acceptor site at position c.1951+1220 is in green: scores calculated by various algorithms are shown (SpliceSiteFinder-like; MaxEntScan; GeneSplicer). Scores of the cryptic donor splice site at position c.1951+1297 (in blue) are not changed between the wild type and variant alleles, but this site will ultimately be used to produce the 77 bp pseudoexon (displayed as a red horizontal line).

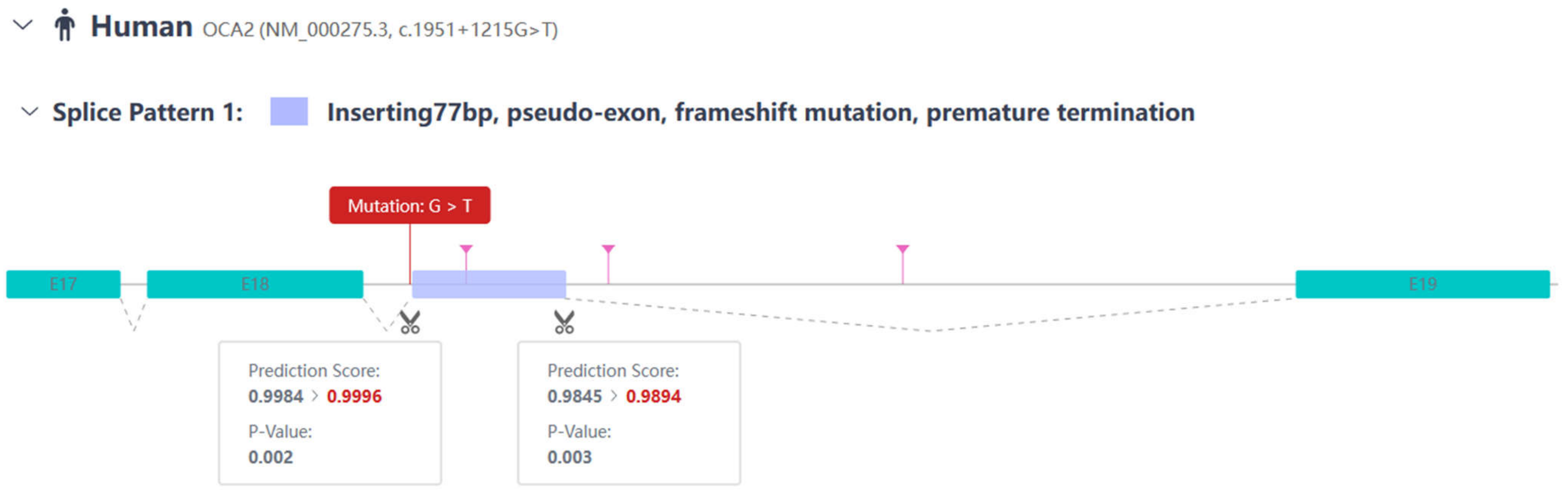


**Supplementary Figure 2B: RNA SPLICER PREDICTION.** RNA Splicer (https://rddc.tsinghua-gd.org/) predicts the activation of a cryptic acceptor splice site and usage of a cryptic donor splice site, thus triggering the insertion of a 77 bp pseudo exon (violet box) in intron 18.

a)

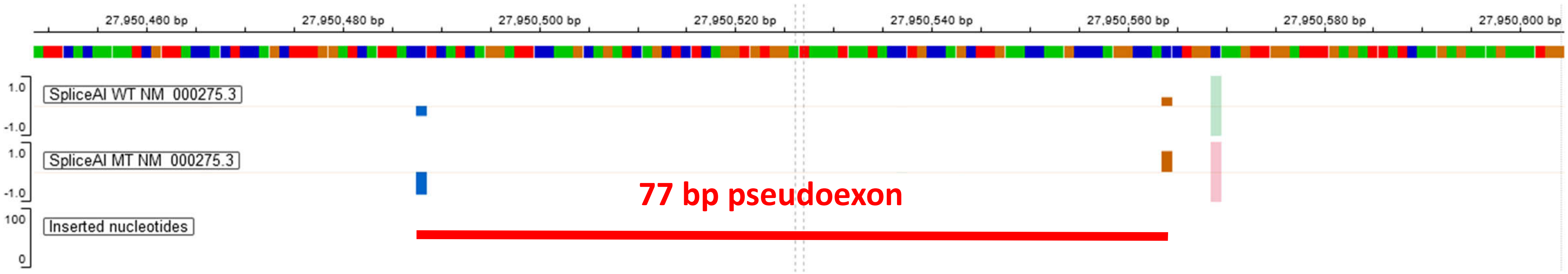


b)

| | | |
|---|---|---|
| spliceAI lookup (500) AG: | 0.40 (-5) | *Acceptor Gain, Δ score 0-1 (relative position in bp), thresholds ≥ 0.2\|0.5\|0.8 for impact* |
| spliceAI lookup (500) AL: | 0.01 (40) | *Acceptor Loss, Δ score 0-1 (relative position in bp), thresholds ≥ 0.2\|0.5\|0.8 for impact* |
| spliceAI lookup (500) DG: | 0.42 (-81) | *Donor Gain, Δ score 0-1 (relative position in bp), thresholds ≥ 0.2\|0.5\|0.8 for impact* |
| spliceAI lookup (500) DL: | 0.00 (51) | *Donor Loss, Δ score 0-1 (relative position in bp), thresholds ≥ 0.2\|0.5\|0.8 for impact* |

**Supplementary Figure 2C: SPLICE AI PREDICTIONS.**

a) Splice AI Visual (integrated in the Mobidetails variant interpretation tool (https://mobidetails.iurc.montp.inserm.fr/MD): green and pink vertical bars show the variation site on the wild-type and mutant track, respectively. Positive orange bars represent the predicted cryptic acceptor site which is activated by the NM_000275.3:c.1951+1215G>T variant; the negative blue bars represent the cryptic donor site which is activated. The 77 bp pseudoexon is displayed as a red horizontal line.

b) SpliceAI prediction scores show that a splice acceptor site is activated 5 bp downstream of the variant, and a splice donor site is activated 81 bp downstream of the variant.
