## Supplementary Figure 3 for "GENOTYPIC SPECTRUM OF ALBINISM IN MALI"

### Slide 1
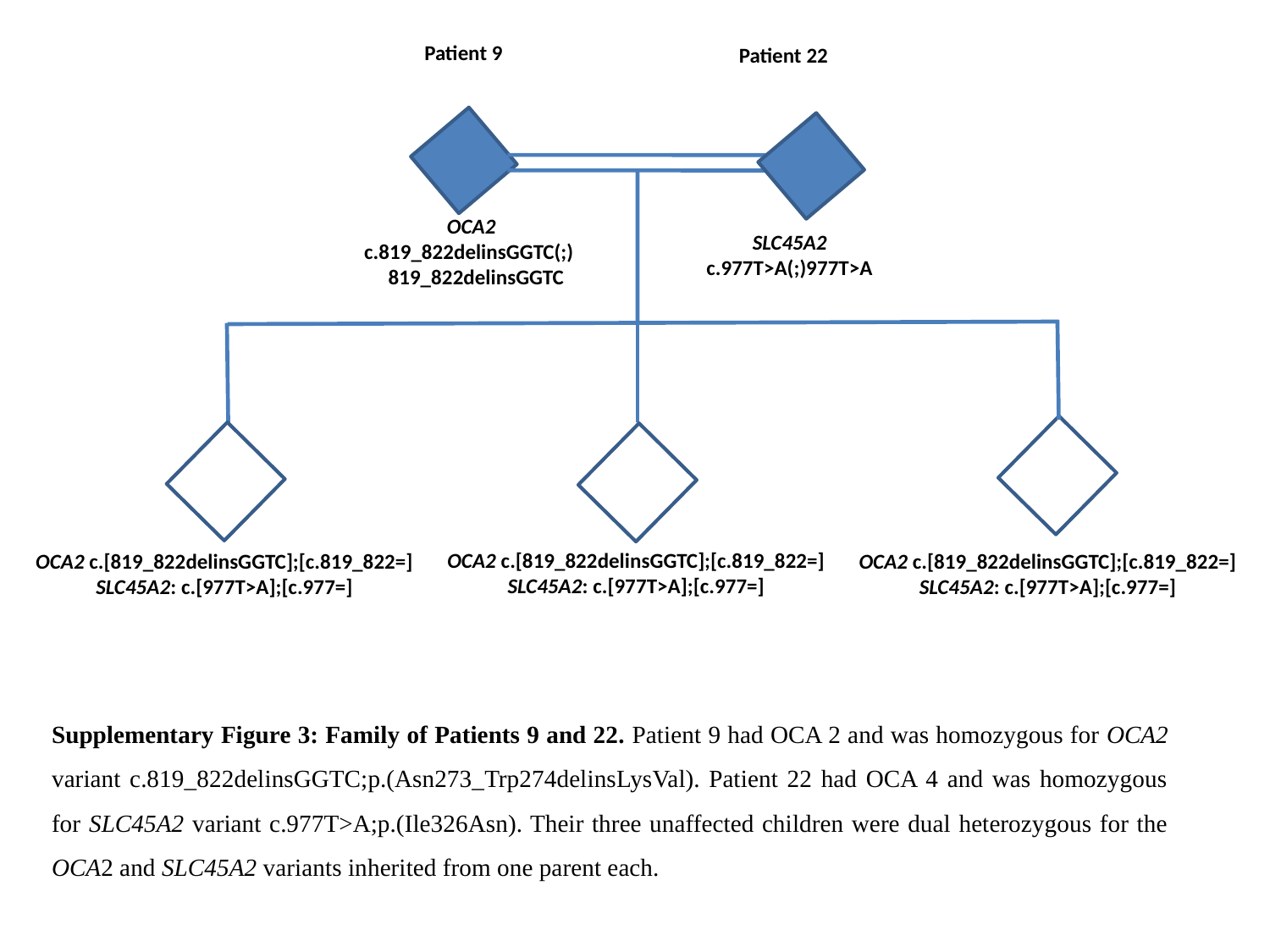

Patient 9
Patient 22
 OCA2
c.819_822delinsGGTC(;)
 819_822delinsGGTC
SLC45A2
c.977T>A(;)977T>A
 OCA2 c.[819_822delinsGGTC];[c.819_822=]
 SLC45A2: c.[977T>A];[c.977=]
 OCA2 c.[819_822delinsGGTC];[c.819_822=]
 SLC45A2: c.[977T>A];[c.977=]
 OCA2 c.[819_822delinsGGTC];[c.819_822=]
 SLC45A2: c.[977T>A];[c.977=]
Supplementary Figure 3: Family of Patients 9 and 22. Patient 9 had OCA 2 and was homozygous for OCA2 variant c.819_822delinsGGTC;p.(Asn273_Trp274delinsLysVal). Patient 22 had OCA 4 and was homozygous for SLC45A2 variant c.977T>A;p.(Ile326Asn). Their three unaffected children were dual heterozygous for the OCA2 and SLC45A2 variants inherited from one parent each.
